# A systematic review and meta-analysis protocol to compare the risk of attrition among prenatal vitamin D supplemented gestational diabetes mellitus patients with its non-recipients

**DOI:** 10.1101/2020.05.22.20110262

**Authors:** Sumanta Saha

## Abstract

**Aim:** Recently, several clinical trials have tested the effect of prenatal vitamin D supplementation in gestational diabetes mellitus (GDM) patients and their newborns. However, their participant attrition, an important determinant of the internal validity, remains poorly explored. Therefore, this review protocol is proposed, which aims to compare it between prenatal vitamin D supplemented and not supplemented GDM patients.

**Method:** Randomized clinical trials studying the above will be searched mainly in different electronic databases. It will search for the English language articles irrespective of publication date. Data concerning the reviewed trials’ design, population, interventions, and outcome of interest will be extracted. Subsequently, their risk of bias will be assessed. Outcome data between the interventions will be juxtaposed meta-analytically to estimate the risk ratio. Next, the statistical heterogeneity (by Chi^2^ and I^2^ statistics) and publication bias (using funnel plot and Eggers test) among the trials will be evaluated. Subgroup analysis will follow if the heterogeneity is high. Finally, a sensitivity analysis will iterate the meta-analysis using an alternative model and by eliminating a trial every time. Statistical significance will be determined at p<0.05 and 95% CI. All analyses will be done in Stata. If a meta-analysis is not possible, a narrative reporting will ensue.

**Results:** The review will follow the PRISMA reporting guideline. Any statistically significant finding’s evidence quality will be graded by the GRADE approach.

**Conclusion:** The proposed review will compare the risk of loss to follow up among GDM patients complemented and not complemented with prenatal vitamin D.

## INTRODUCTION

GDM is a medical complication of pregnancy.[1] It manifests as glucose intolerance (of any degree) that is detected or developed for the first time during pregnancy.[2] With the rising trend in obesity and a sedentary lifestyle, its prevalence in reproductive age group females is increasing globally.[3] A 50 gm and 1-hour glucose challenge test is used as a screening test to diagnose GDM between 24-28 weeks of gestation.[2] GDM’s complications can affect both the mother and her neonate. Maternal complications may include an increased risk of type 2 diabetes development, cesarean section, pre-eclampsia, and polyhydramnios.[2,4,5] Some of the fetal complications include newborn hypoglycemia, hyperbilirubinemia, and increased perinatal mortality.[2] GDM is classified as A1GDM if managed primarily with dietary modifications and A2GDM when medication is also required for the treatment.[2] Besides, compliance with self-monitoring of blood glucose levels is also an essential component of GDM management.[1,6] The dietary advice, blood glucose monitoring, and the use of insulin on a required basis help to decrease the perinatal complications.[7] The medical nutrition therapy in GDM primarily helps to maintain optimal glycemic control by regulating the caloric intake.[6] While dietary intervention is a vital treatment component of both A1GDM and A2GDM,[2] the role of supplements in GDM mothers is not well established. In this regard, vitamin D has drawn significant attention to the research community as a novel supplement.

### Vitamin D in GDM

Evidence suggests that GDM patients tend to be vitamin D deficient.[8–11] Besides, due to the rapid development of the fetus (e.g., bone calcification), pregnancy itself increases the risk of vitamin D deficiency, especially during the last stages of pregnancy.[12] Therefore, it’s sensible to conduct trials to test the effects of vitamin D supplementation in GDM patients. Such supplementation is further justified by the fact that the fetus depends on the mother for vitamin D.[12] Recently, several trials have studied the effect of antenatal vitamin D supplementation in GDM mothers. A meta-analysis of such trials found that certain blood glucose and lipid parameters might improve (HOMA-IR, QUICKI, and LDL-cholesterol) while others (affect FPG, insulin, HbA1c, triglycerides, total- and HDL-cholesterol levels) might not change.[13] Some of the trials comparing the effect of vitamin D supplementation on gestational weight and BMI of GDM patients did not find any statistically significant difference with the placebo.[14–16] Certain trials have explored the perinatal complications of the GDM mothers and their neonates, in vitamin D supplemented GDM mothers. In these trials, incidence of cesarean section,[17,18]preterm delivery,[17–19] pre-eclampsia,[17–19] hyperbilirubinemia[17,18]and hypoglycemia[18,19] did not vary between vitamin D receiving and non-receiving group. In one of these trials, vitamin D supplementation decreased the cesarean section, hyperbilirubinemia, and hypoglycemia.[19]

### Incomplete outcome data and vitamin D supplementation

Amidst of these growing numbers of trials, it is crucial to study the validity of their findings due to the missing outcome data. Such bias can also creep in the meta-analysis that extracts data from these trials. Notably, attrition is common even in the adequately conducted RCTs.[20] In vitamin D supplemented trials on GDM patients, both the continuous (e.g., gestational weight, body mass index, blood glucose and lipid parameters) and dichotomous (e.g., the frequency of CS, pre-eclampsia, preterm delivery, neonatal jaundice, and newborn hypoglycemia) outcomes can be affected due to missingness of participants from the trials. For dichotomous outcomes, comparison of the missing data to the frequency of events ratio across multiple trials helps to juxtapose their risk of bias.[21] Whereas, for continuous outcomes, the proportion of missing data determines the risk of bias. Among two hypothetical trials with identical observed mean and missing values’ average, the mean difference of the outcomes between the compared intervention groups will be higher in the trial with a higher number of participant attrition.[21]

Besides the above, the causes of missingness between the intervention arms also determine the risk of bias in RCTs. For instance, in two trials investigating the effects of antenatal vitamin D supplementation on GDM patients, the reasons for missingness weren’t identical between the treatment groups. While intra-uterine fetal demise, placental abruption, and hospitalization were the causes of missing outcome data in the vitamin D recipients, insulin therapy, pre-eclampsia, hospitalization, and placental abruption were the causes of that in the comparison group.[15,16] Along with this, to minimize the risk of bias, a balance in the frequency of attrition between the intervention groups of a trial is required.[21]

Given these implications of incomplete outcome data, it is imperative to investigate how it differs between prenatal vitamin D supplemented and not supplemented GDM patients.

### Intervention description

Vitamin D is a fat-soluble hormone, available from diet and supplements in two inactive forms: D2 (ergocalciferol) and D3 (cholecalciferol).[22–24] The D3 form is additionally produced in the skin on exposure to the sun.[22,23] Upon hydroxylation of pre-vitamin D in the liver, the main circulatory form of vitamin D (bound to albumin or in a free state), 25-hydroxyvitamin D (25(OH)D) is produced.[25] Then, this 25(OH)D is converted into its active form, calcitriol (1,25(OH)2D).[22,26] Calcitriol, stimulates intestinal calcium and phosphorus absorption and renal reabsorption of calcium via its receptors.[23] It also plays its physiologic role in pregnancy on binding to the vitamin D receptors in the uteroplacental tissue.[22,26] The recommended dietary allowance and the tolerable upper level of intake of vitamin D in pregnancy are 600 and 4000IU, respectively.[22]

The trials on GDM patients have tested vitamin D in different dosages. As an oral supplement, some trials have prescribed 50,000 IU of vitamin D at two to three weeks interval for about three to eight weeks.[15,16,27,28] Trials in which participants were advised to take it orally every day, recommended at a dose of 200-500IU.[29,30] Use of its injectable (intramuscular) forms was at a dosage of 300,000 IU.[12] In RCTs on GDM patients, the antenatal supplementation of vitamin D has been tested as a sole[12,27,29] or co-supplement (with other micronutrients like zinc, calcium, and magnesium).[16,31]

### Why this proposed systematic review and meta-analysis is required?

Parallel to the rising number of trials testing vitamin D supplementation’s effect in GDM patients, systematic review and meta-analysis using these trials are also increasing. Presently, such reviews have primarily studied the influence on glucose hemostasis parameters and lipid profiles.[13,32–34] Besides, a protocol for systematic review and meta-analysis exists that intends to determine how the intervention might influence the various maternal health parameters like body mass index, gestational weight, and 25(OH)D levels.[24] However, best known to this author, the risk of attrition in these trials has not been systematically explored. Therefore, to study this poorly explored area of the GDM literature, this systematic review protocol is proposed here.

The review aims to compare how the risk of loss to follow up in the existing RCTs vary between prenatal vitamin D supplemented and not supplemented GDM patients.

## METHODS

Studies matching the following eligibility criteria will be included in the proposed review. Inclusion criteria: 1. Study design: Parallel arm (any number of arms) randomized controlled trials, irrespective of their blindness and duration. 2. Study population: Women of any age diagnosed with GDM during their concurrent pregnancy. Women will be recruited irrespective of their gravida, parity, or previous history of GDM diagnosis. 3. Intervention arm: A trial must test vitamin D as a sole or co-supplement with other nutrients in one or more of its treatment arms. 4. Comparator arm: The comparator arm can receive any supplement that doesn’t contain vitamin D or placebo or no intervention. 5. Outcome: Attrition in at least one of the treatment arms post-randomization is required for inclusion in the review. The trialists’ exclusion of available outcome data from the statistical analysis will not form the inclusion criteria of the review.

GDM diagnosis and its management and the dosage, regimen, and route of administration of the interventions (in each of the treatment arms) will be accepted as per the trialists.

Exclusion criteria: 1. Any study design besides the above, like crossover trial or quasi-experimental study or observational study. 2. Women diagnosed with diabetes of non-GDM variants like diabetes type 1 or type 2.

This review protocol has been submitted to the PROSPERO for registration.

### Database search

Electronic databases (PubMed, Embase, and Scopus) will be searched for the title and abstract of literature published in the English language with no limit to date. A draft of the search strategy to be used in PubMed is presented here: "vitamin D" OR calciferol OR “vitamin D2” OR ergocalciferol OR “vitamin D3” OR cholecalciferol AND “gestational diabetes” OR GDM. Subsequent MeSH terms will be used in this search “cholecalciferol,” “ergocalciferols,” and “diabetes, gestational.” The filters “Clinical Trial” and “Randomized Controlled Trial” will be used to narrow down the search results to RCTs. Additional searches will be done in the references of the reviewed trials.

### Study selection

The search output from the respective electronic databases will be uploaded into a systematic review software, Rayyan.[35] Then, the remaining papers will be scanned by the authors, after eliminating the duplicates. On reading the title and abstract, if a paper seems to meet the eligibility criteria mentioned above, it will be read in entirety. Besides, a full-text reading will ensue when a paper can’t be excluded decisively.

A record of the last date of the literature search and the reasons for eliminating papers read in the full text will be retained. The trial with an overall higher number of missing outcome data will be reviewed when multiple (eligible) trials source data from the same trial population.

### Data extraction

Data of the study design, population characteristics, compared interventions, and outcome of interest will be abstracted from each of the reviewed trials. In the study design, information on randomization, blinding, trial registration number, number of intervention arms, single or multi-centeredness, trial duration, nation where the trial was conducted, ethical clearance, participant consent, and funding information will be extracted. Subsequent participant details will be captured- the number of participants randomized to each of the intervention arms, their mean age, their diagnosis, and the criteria (e.g., American Diabetic Association criteria)[36,37] used to make the diagnosis. Next, concerning the interventions, the types of intervention used in respective treatment arms along with their dosage, regimen, and mode of administration will be collected. Finally, for the outcome of interest, the number of participants missing from each of the treatment arms will be recorded with their reason for drop out of the trial.

### Risk of bias assessment

The Cochrane collaboration tool will be used to assess the selection, performance, detection, attrition, reporting, and other bias. For respective trials, each of these biases will be evaluated and categorized as low, high, or unclear.[21] The method of random sequence generation and its mode of allocation to keep participants blinded about the intervention will be used to decide on the selection bias. Blinding information of study personnel and participants and that of outcome assessors will be utilized to judge the performance and detection bias, respectively. Then, using the balance and reasons for participant attrition from the intervention groups, the attrition bias will be assessed. Next, for the reporting bias, the trial results will be juxtaposed to its pre-specified intentions. Finally, any other bias not categorizable into any of the above types, will be included as a miscellaneous bias.

### Author role

SS^1^ will perform the electronic database search. The review authors will independently select the eligible trials, abstract their data, and assess their risk of bias. A third-party help will be sought when authors fail to resolve any disagreement by discourse.

### Meta-analysis

Since the missing outcome data is dichotomous, the risk ratio will be estimated by a meta-analytic comparison between antenatal vitamin D supplemented and not supplemented intervention groups. The model for meta-analysis will not be based on predetermined statistical inconsistency, rather, guided by the clinical heterogeneity. The use of a random-effect model (DerSimonian and Laird method) or fixed-effect model (inverse-variance method) will be based on the clinical heterogeneity or homogeneity of the trials, respectively. For an unbiased summary estimate, trials at a high risk of bias will be excluded from the meta-analysis. Its statistical significance will be determined at p<0.05 and 95% confidence interval. If the estimated risk of attrition is statistically significantly different between the contrasted treatment arms, it will be graded applying the GRADE approach (by GRADE Working Group (2004)).[38]

### Special situations in meta-analysis

Here, anticipated hurdles to meta-analysis are explored. If a trial has attrition in any one of its treatment arms, 0.5 will be added to each cell of the 2×2 table as continuity correction. Another situation may include the testing of an intervention (e.g., vitamin D supplementation) in several treatment arms of a trial; outcome data will be combined across these intervention groups.

### Heterogeneity and publication bias

By utilizing the I^2^ and Chi^2^ statistics, the statistical heterogeneity will be assessed. I^2^ values of 25, 50, and 75%, will be categorized as low, moderate, and high heterogeneity.[39] Chi^2^ statistics will determine the heterogeneity at p<0.1.[21]An assessment of a high statistical inconsistency will be done by subgroup analysis. Based on the clinical characteristics, the subgroup analysis will be done. For example, it may be based on GDM treatment (between A1GDM and A2GDM) or latitude (within nations with ample and little sunlight exposure) or the mode of vitamin D use (among its use as a sole and co-supplement).

The publication bias will be determined by funnel plots and contour enhanced funnel plots. Eggers test will also be used for the same when ten or more trials are available for the meta-analysis.

### Sensitivity analysis

The robustness of the meta-analysis will be evaluated by repeating the meta-analysis with an alternative model (random-effect or fixed-effect). Additional iteration of the meta-analysis will involve the elimination of a trial each time using both the random-effect and fixed-effect models.

All analyses will be done in Stata statistical software, version 16 (StataCorp, College Station, Texas, USA). If quantitative analysis is not possible, a narrative reporting will follow. The reporting of the review will adhere to Preferred Reporting Items for Systematic reviews and Meta-Analyses guidelines (PRISMA).[40]

## DISCUSSION

To summarize, several trials have contemporarily tested the effects of antenatal vitamin D supplementation on GDM mothers and their infants; however, it’s not clear if missing outcome data threatens the internal validity of these trials and the meta-analyses that depend on these. Therefore, to study how such intervention determines the risk of loss to follow up, this systematic review and meta-analysis protocol is proposed. The trials meeting the inclusion criteria will be searched in different electronic databases. Then, their data of study design, participants, interventions compared, and the outcome of interest will be abstracted, and the risk of bias will be assessed. If comparable outcome data are available, they will be compared meta-analytically; otherwise, a narrative review will follow.

Next, discussed are the planned review’s possible implications and strengths. It will provide future trialists with greater insight on participants missingness in trials testing the effect of antenatal vitamin D supplementation in GDM patients, which will consequently address the attrition problem better and help the clinical trials to retain their internal validity. Simultaneously, it will aid in evaluating the robustness of meta-analyses depending on such trials. Furthermore, this will perhaps be the first systematic review to explore the context; hence, the generated evidence will be novel. Besides, as the review will be based on RCTs, the highest level of the epidemiologic evidence, the findings are expected to be rigorous. Additionally, not restricting the database search to any date limit will allow the literature search to be exhaustive.

Despite these strengths, certain limitations are anticipated. If most of the reviewed trials use vitamin D as a co-supplement with some other nutrient, due to the flexible inclusion criteria, it might be difficult to distinguish the vitamin D’s effect. Likewise, accepting the GDM treatment as per the trialists may cause recruitment of a substantial number of trials using insulin and/or oral hypoglycemics to treat GDM. In such situation, it may become difficult to separate its effect from that of the vitamin D supplements.

Lastly, the inclusion of articles published in the English language only will decrease the possibility of identifying relevant trials (if any) published in any other language.

## CONCLUSION

The proposed review will compare the risk of participants’ attrition between GDM patients receiving and not receiving vitamin D supplementation antenatally.

## Data Availability

not applicable

